# Genomic variation associated with cardiovascular disease progression following preeclampsia: A systematic review

**DOI:** 10.1101/2023.05.02.23289430

**Authors:** Gayathry Krishnamurthy, Phuong Tram Nguyen, Bao Ngoc Tran, Hoang T Phan, Shaun P. Brennecke, Eric K Moses, Phillip E. Melton

## Abstract

**Background:** Women with a history of preeclampsia (PE) have been shown to have up to five times the risk of developing later-life cardiovascular disease (CVD). While PE and CVD are known to share clinical and molecular characteristics, there are limited studies investigating their shared genomics (genetics, epigenetics or transcriptomics) variation over time. Therefore, we sought to systematically review the literature to identify longitudinal studies focused on the genomic progression to CVD following PE.

**Methods:** A literature search of primary sources through PubMed, Scopus, Web of Science and Embase via OVID was performed. Studies published from January 1^st^, 1980, to February 02^nd^, 2023, that investigated genomics in PE and CVD were eligible for inclusion. Studies that did not include CVD or related risk factors as outcome, were in non-human species or focused on pregnancy complications other than PE were excluded. Included studies were screened based on Cochrane systematic review guidelines in conjunction with the PRISMA 2020 checklist. Eligible articles were further assessed for quality using the Newcastle-Ottawa scale.

**Results:** A total of 8929 articles were screened with 14 studies subjected to quality assessment. Following further evaluation, six studies were included for final review. All six of these studies were heterogenous in regard to CVD/risk factor as outcome, gene mapping approach, and in different targeted genes. The only common variable across all six studies was use of a case-control study design.

**Conclusions:** Our results provide critical insight into the heterogeneous nature of genomic studies investigating CVD following PE and highlight the urgent need for longitudinal studies to further investigate the genetic variation underlying the progression to CVD following PE.

## INTRODUCTION

Cardiovascular disease (CVD) is the global leading cause of morbidity and mortality in women^(1)^. Recent studies have shown biological sex disparities in the CVD pathophysiology, clinical diagnosis and responsiveness to management^(2)^. Dyslipidaemia, hypertension, smoking, obesity, and diabetes are some of the major CVD risk factors common between men and women^(3)^. In addition, women can present with additional medical concerns which make CVD more challenging to identify^(1, 4)^. Pregnancy complications are now considered an important later-life CVD risk factor in women^(5, 6)^. This is especially apparent in women who have had the hypertensive disorder of pregnancy, preeclampsia (PE). PE is characterised by new-onset hypertension and proteinuria or other organ dysfunction after 20 weeks of gestation^(7)^. PE, which affects 2% to 8% of pregnancies worldwide, can cause disruptions to the maternal endothelium, leading to a decrease in angiogenesis and reduced blood flow to organs and tissues^(8, 9)^. Women with a history of PE have been shown to have up to five times the risk of developing later-life CVD when compared to their normotensive counterparts^(10)^.

PE and CVD are known to share molecular pathological features, including endothelial dysfunction, metabolic abnormalities, inflammatory response, oxidative stress and hypercoagulability^(11, 12)^. For example, increased levels of systemic inflammation in PE have been linked to a higher risk of atherosclerosis, endothelial dysfunction and early onset of arterial stiffness^(13)^. Moreover, the adipocyte-derived hormone leptin, a marker of increased CVD and obesity risk, has also been found to be elevated in women with PE^(12)^. Maternal pre-pregnancy BMI is strongly associated with increased risk of PE^(14, 15)^ and elevated BMI is a hallmark risk factor for increased CVD risk.

To model the complex biological relationship between CVD and PE, we and others have undertaken several studies using a wide array of gene-mapping techniques^(16–19)^. These range from candidate gene studies to whole-genome sequencing. These studies seek to identify specific loci within the human genome that are associated with either a PE or CVD-specific outcomes/traits that may underly both conditions. A common approach in these genetic studies is to look for overlap between CVD and PE using the concept of pleiotropy, which refers to when a single gene or loci influences two seemingly unrelated traits. For example, our genetic dissection of the PE susceptibility loci on chromosome 2q22 identified variants in four genes (*LCT, LRP1B, GCA*, *RND3*) that were associated with PE in an Australian family cohort^(19)^. These variants were also associated with cardio-metabolic traits in both the San Antonio Family Heart Study and Australian adolescents from the Raine Study^(18)^. While these previous genetic studies provide important information on the shared genetic susceptibility loci for PE and CVD they do not inform on specific genes or loci that are involved in the progression from PE to later-life CVD.

In addition to above referenced genetic studies, there are also several studies that used an array of recent genomic technologies to investigate the biological relationship between PE and CVD. These studies focus on gene regulation (epigenetics) and gene expression (transcriptomics). For example, a meta-analysis investigated differential expression of PE in placental tissue and whole blood from CVD patients and observed 22 genes common to both PE and CVD, from 925 PE and 181 CVD differentially expressed genes. This study also identified common biological pathways including oxidative stress, interleukin signalling, inflammation-mediated chemokines and cytokines that are known to play a role in the complex pathogenesis of both disorders^(20–22)^. Other studies have focused on differential DNA methylation and miRNAs (microRNAs) which are known to influence both PE and CVD but there have been limited efforts to investigate the progression of these genomic loci from PE to later-life CVD in women.

The aim of our study was to systematically review the literature to identify genomic loci involved in the progression to CVD following PE in the same women over time. Findings from this review may inform more impactful research strategies for identifying genomic loci influencing increased CVD risk following PE.

## METHODS

An examination of primary research literature was performed to identify genomic variation associated with the progression to CVD following PE. The Cochrane handbook for systematic reviews of interventions (2nd edition) was followed^(23)^ along with the Preferred Reporting Items for Systematic Reviews and Meta-Analyses (PRISMA) 2020 statement (Supplementary file 1) to enhance the study protocol and ensure comprehensive reporting of findings^(24)^.

### Information Sources and Search Strategies

The primary literature search was conducted on August 27^th^, 2021, and an updated search on February 2^nd^, 2023, using four electronic databases: PubMed, Scopus, Web of Science and Embase via OVID. Databases were searched using the title, abstract and full-text fields to find all significant articles. The search keywords used included “cardiovascular disease”, “heart arrhythmias”, “stroke”, “cardiometabolic risk”, “hypertension”, “dyslipidaemia”, “cardiomyopathy”, “atherosclerotic heart diseases”, “rheumatic heart disease”, “cerebrovascular disease”, “coronary artery disease”, “heart failure”, “heart valve disease”, “ischaemic heart disease”, “inflammatory heart disease”, “heart disorder”, “cardiac arrest”, “hypertensive heart disease”, “carditis”, “peripheral artery disease”, “myocardial infarction”, “acute coronary syndrome”, “cardiac failure”, “left ventricular systolic dysfunction”, “preeclampsia”, “toxaemia”, “maternal syndrome”, “pregnancy complication”, “pregnancy-specific disorder”, “pregnancy-induced hypertension”, “maternal hypertension”, “eclampsia”, “HELPP syndrome”, “genetic”, “candidate gene studies”, “association analysis”, “linkage studies”, “Genome-wide association studies”, “Genome-wide linkage studies”, “gene-gene interactions”, “gene-environmental interactions”, “epistasis”, “heritability”, “DNA methylation”, “epigenetics”, “microRNA”, “histone modification”, “chromatin modification”, “epigenetic modification”, “Epigenome-wide association studies”, “posttranslational regulation”, “transcriptional gene silencing”, “nucleosome remodelling”, “non-coding RNA regulation” and “RNA editing”. Detailed search keywords are mentioned in the data Supplementary files 2 and 3.

### Inclusion and Exclusion Criteria

This systematic review included original articles published in English from-1980 until February 2^nd^, 2023, in case-control, cohort, or cross-sectional study designs. We chose post-1980s due to significant advancements in genomic research and mapping techniques. Publications relevant to progression of PE to CVD endpoints (e.g., coronary artery disease, stroke, etc) or CVD risk factors (e.g., systolic blood pressure, cholesterol level) in women based on genetic, epigenetic or transcriptomic factors were included. PE was defined either based on the clinical outcome presented during pregnancy or as a history of PE from self-reported questionnaires answered by the participants. Traditionally, PE is clinically defined by a presence of hypertension (≥ 140/90 mmHg) along with proteinuria (≥ 0.3 g/24 h or a dipstick reading of ≥1+) when measured at least twice after 20 weeks of pregnancy^(25)^. The self-reported responses obtained through questionnaires were crosschecked with the medical records of the participants by an Obstetrician/Gynaecologist to ensure the accuracy of the data.

The articles were restricted to humans and peer-reviewed empirical studies. Studies were excluded if: (i) CVD was not evaluated as the outcome; (ii) they were on behavioural CVD risk factors, including diet, physical activity, alcohol consumption, and tobacco use; (iii) they were solely aimed at other pregnancy complications other than PE, such as gestational hypertension, gestational diabetes, stillbirth, small-for-gestational-age; (iv) focused on offspring rather than mothers; (v) based on animal models; (vi) there was a lack of genetic, epigenetic or transcriptomic evidence; (vii) they were systematic reviews, discussion papers, case reports, case series, editorials, or conference abstracts; (viii) focused on a topic not related to CVD or PE; and (ix) different women or different cohorts of CVD and PE were used to study the association between both diseases.

### Data Screening, Selection and Extraction

The research articles from all four databases were imported to EndNote 20 to remove duplicates^(26)^. The remaining articles were exported to Rayyan Systematic Review software for further assessment, screening, selection and extraction of data^(27)^. Three reviewers (G.K., T.N., and N.T.) collaborated to screen and select relevant studies. First, reviewers independently conducted a blind assessment of the titles and abstracts of all studies. The eligible articles were then subjected to full-text screening based on the inclusion and exclusion criteria. Finally, disagreements or inconsistencies between the three reviewers were addressed through discussion and consultation with a fourth reviewer (P.M.). The reasons for exclusion were clearly stated at the end of each screening stage.

Data were retrieved from the included studies and recorded on a data extraction table. The significant extracted characteristics included study designs, gene identification approaches, number of variants from genes, polymorphisms or miRNAs, participant demographic data, CVD follow-up time after first index pregnancy, and the definition of PE and CVD outcome measures.

### Risk of Bias and Methodological Quality Assessment

The quality level of each study that reached the final screening stage was independently reviewed by three reviewers (G.K., T.N., and N.T.). The evaluation was conducted using the Newcastle-Ottawa Quality Assessment Scale for case-control, cohort and cross-sectional studies^(28)^. The articles were ranked on a scale of 0 to 10 and have been further customised for this review (See Supplementary file 4). The quality assessment criteria were based on case selection, comparability between cases and controls and outcome or exposure. Most studies had more than one study population with different study designs. Hence, the quality of each of these cohorts has been individually assessed, as they could not be categorised into one study design. The reasons for excluding studies despite having a good quality score were mentioned in the Quality Assessment Supplementary Tables S4-S6. Any difference of opinion between the three reviewers was resolved through discussion with the fourth reviewer (P.M.).

### Data Analysis

Three reviewers (G.K., T.N., and N.T.) performed the data extraction using Microsoft Word based on the Cochrane handbook for systematic reviews (2nd edition)^(23)^. Included articles were thoroughly read and classified into two study types: CVD endpoints and CVD risk factors. However, the study design, methodologies, results, and the CVD outcomes analysed differed in each included study. Hence, a meta-analysis was not performed due to cross study heterogeneity.

## RESULTS

### Study Selection

The Systematic Review Flowchart provides a detailed overview of the selection process (Figure 1). The primary search generated 12,988 records including the articles from the updated search. A total of 8,929 studies were then subjected to the title/abstract screening after eliminating duplicates (N=4059). Further, 8,713 additional studies were removed after the eligibility criteria were narrowed to be more specific to the study aim, excluding studies that: (i) were not focusing on both PE and CVD; (ii) were on animal studies; (iii) were systematic and literature reviews; (iv) did not evaluate CVD as the outcome; and (v) lacking genomic evidence. This resulted in the full-text screening of 216 studies. Following the full-text screening, 14 articles were included for quality assessment, excluding 202 studies with any of the above reasons. In addition, the articles which studied other hypertensive disorders of pregnancy and not PE, studies on offspring and those which were not original research were also excluded in the full-text screening stage. After further refinement, eight articles were removed for the following reasons: (i) two studies for using different cohorts for CVD and PE^(19, 29)^; (ii) one article for investigating different women with CVD and a history of PE, although from the same cohort^(30)^; (iii) two for making conclusions on the shared risk of PE and CVD using genes predisposed to CVD from another study^(31, 32)^ and (iv) for not testing for CVD risk^(31, 32)^; (v) two for studying CVD risk in women with PE during delivery, but not conducting any follow-up research on CVD risk^(33, 34)^; and (vi) one study for not mentioning the number of women with PE^(18)^ (see Supplementary Tables S1-S6 for details). The included studies consisted of two studies on CVD endpoints following PE^(35, 36)^ and four on CVD risk factors following PE^(37–40)^ (see Supplementary Tables S1-S6). Ultimately, six articles met the study selection criteria.

**Figure 1:**
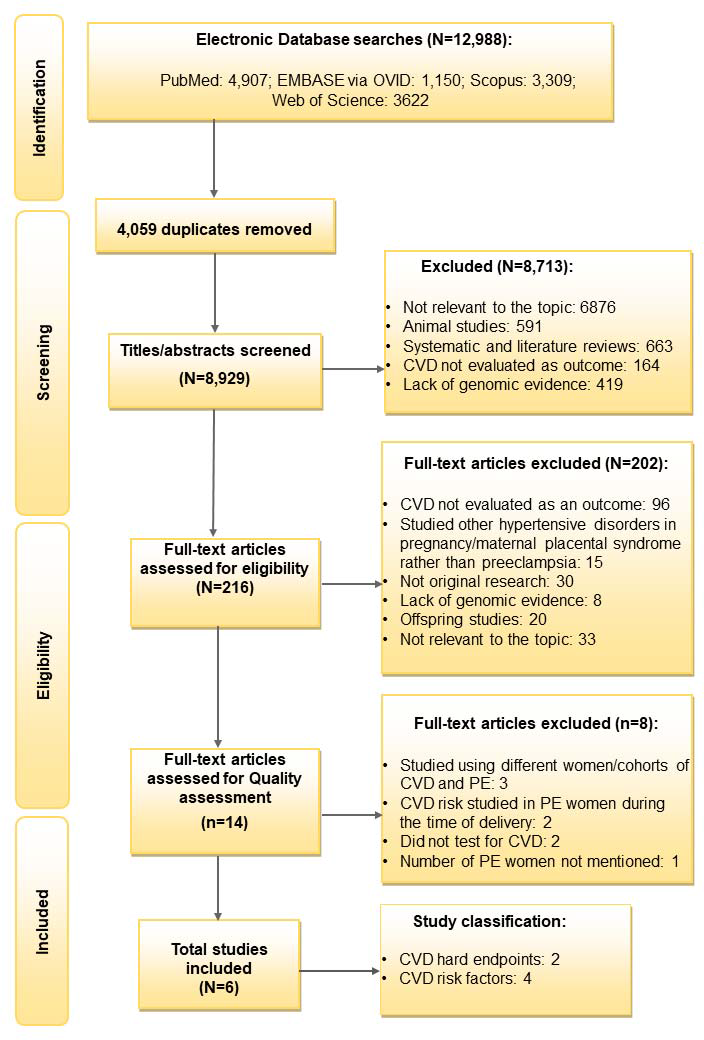
Flowchart showing Study Selection. CVD indicates Cardiovascular disease and PE indicates Preeclampsia

### Quality Assessment

The quality scores of six included studies^(35–40)^ and the excluded eight studies^(18, 19, 29, 30–33, 41)^ that reached the final screening stage are shown in Supplementary Tables S1-S3. The six studies that passed the final screening were all case-control design. Hence, the New-Castle Ottawa scale for case-control studies was used (maximum score: nine); three studies scored 7.5^(36, 37, 40)^, one received 8.5^(35)^ and another obtained 5.5^(39)^. The final included study consisted of two case-control cohorts, and both were assessed for quality separately^(38)^. Among these, cohort 1 scored 6 and cohort 2 scored 6.5^(38)^. Four studies had reliable methods of ascertaining PE from medical records or databases^(35–37, 40)^. However, two studies based their PE diagnosis on self-reported questionnaires^(38, 39)^. All studies investigated a variety of genomic factors involved in the progression to CVD or CVD risk factors following PE in the same women from the same cohorts of PE and CVD.

### Study Characteristics

The study characteristics for the CVD endpoints and CVD risk factors studies are summarised in Tables 1 and 2, respectively. The articles included consisted of three candidate gene studies^(36, 37, 40)^, two miRNA studies^(38, 39)^, and one epigenetic study^(35)^.

**Table 1:**
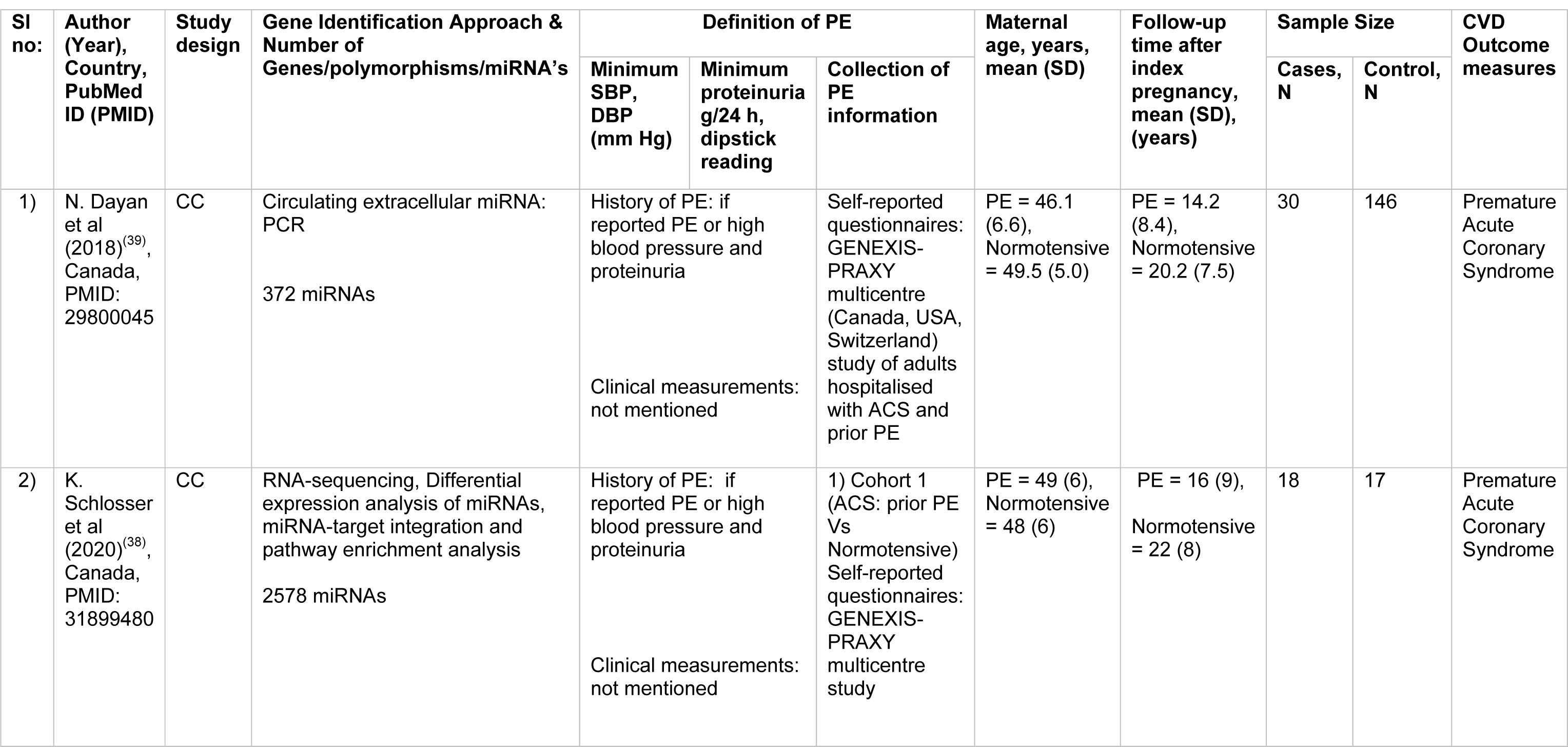
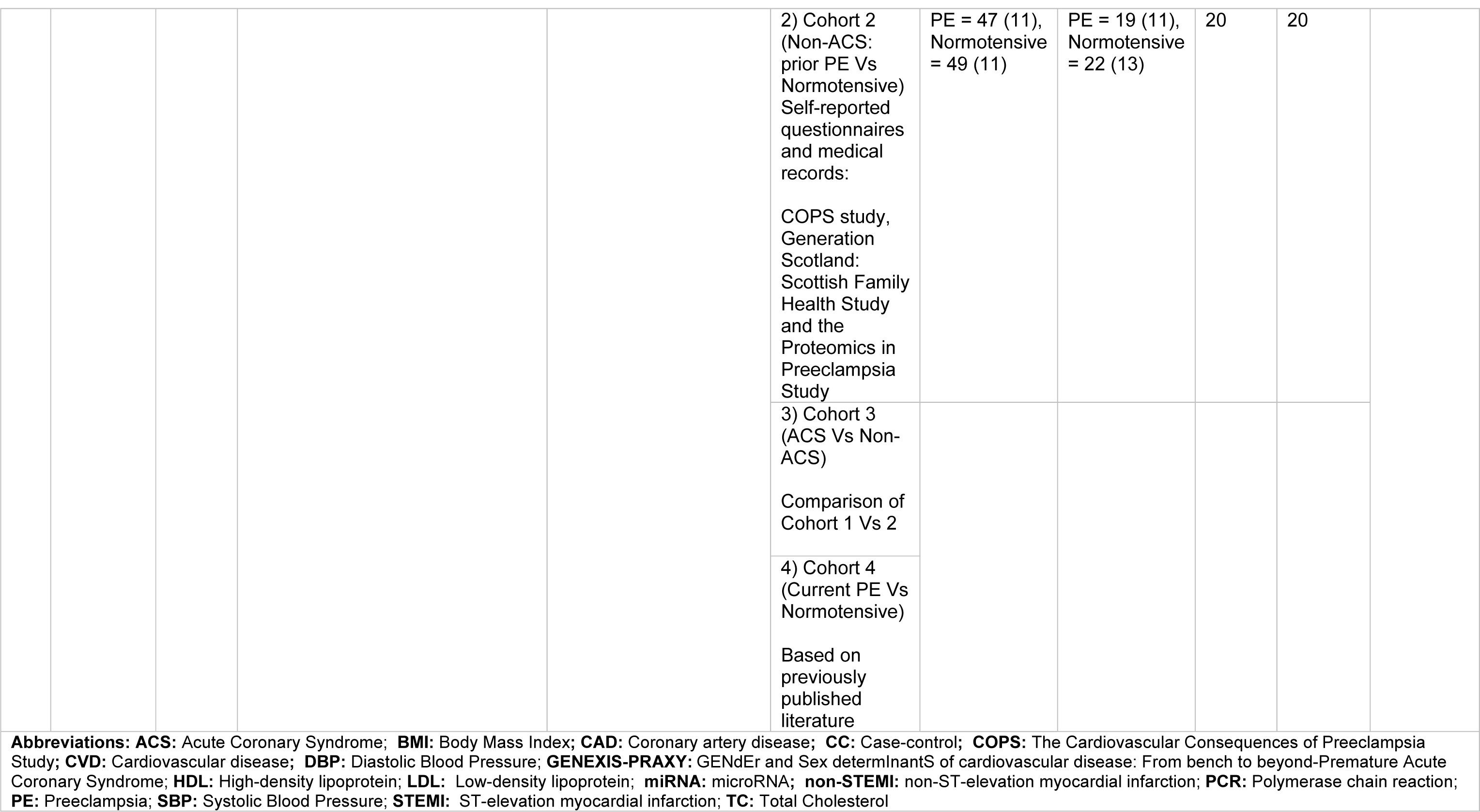
General Characteristics of Studies on PE and CVD hard endpoints.

**Table 2:**
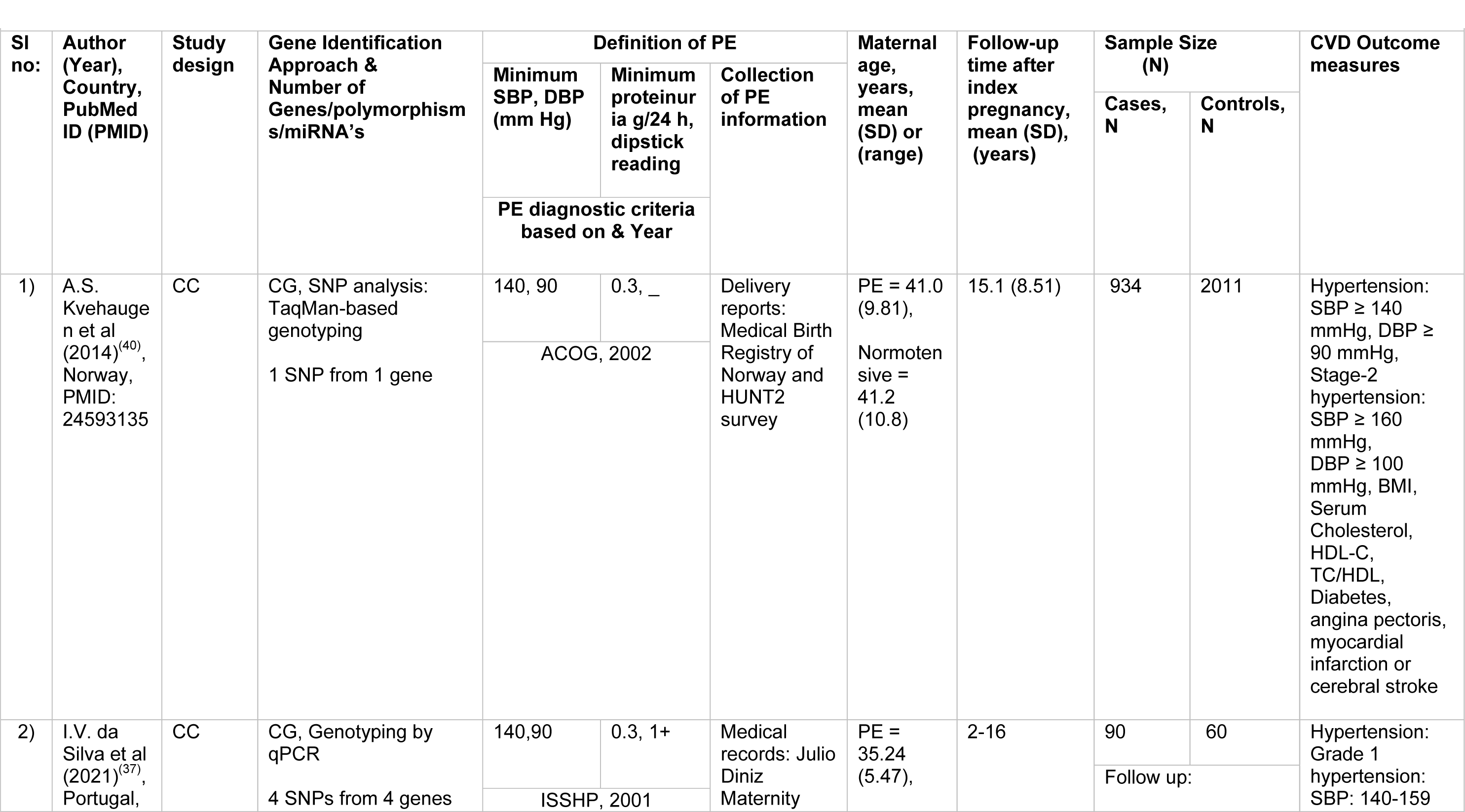
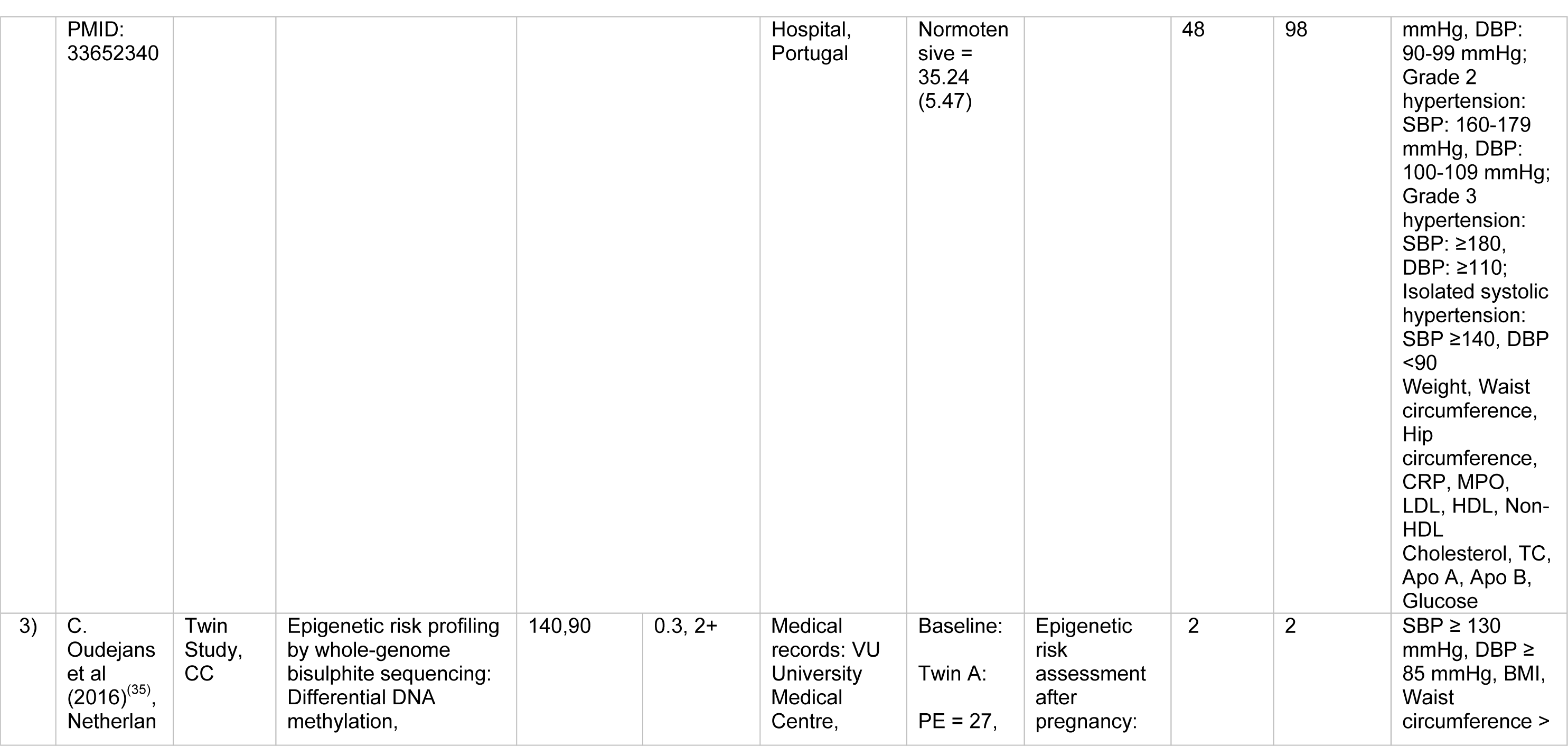
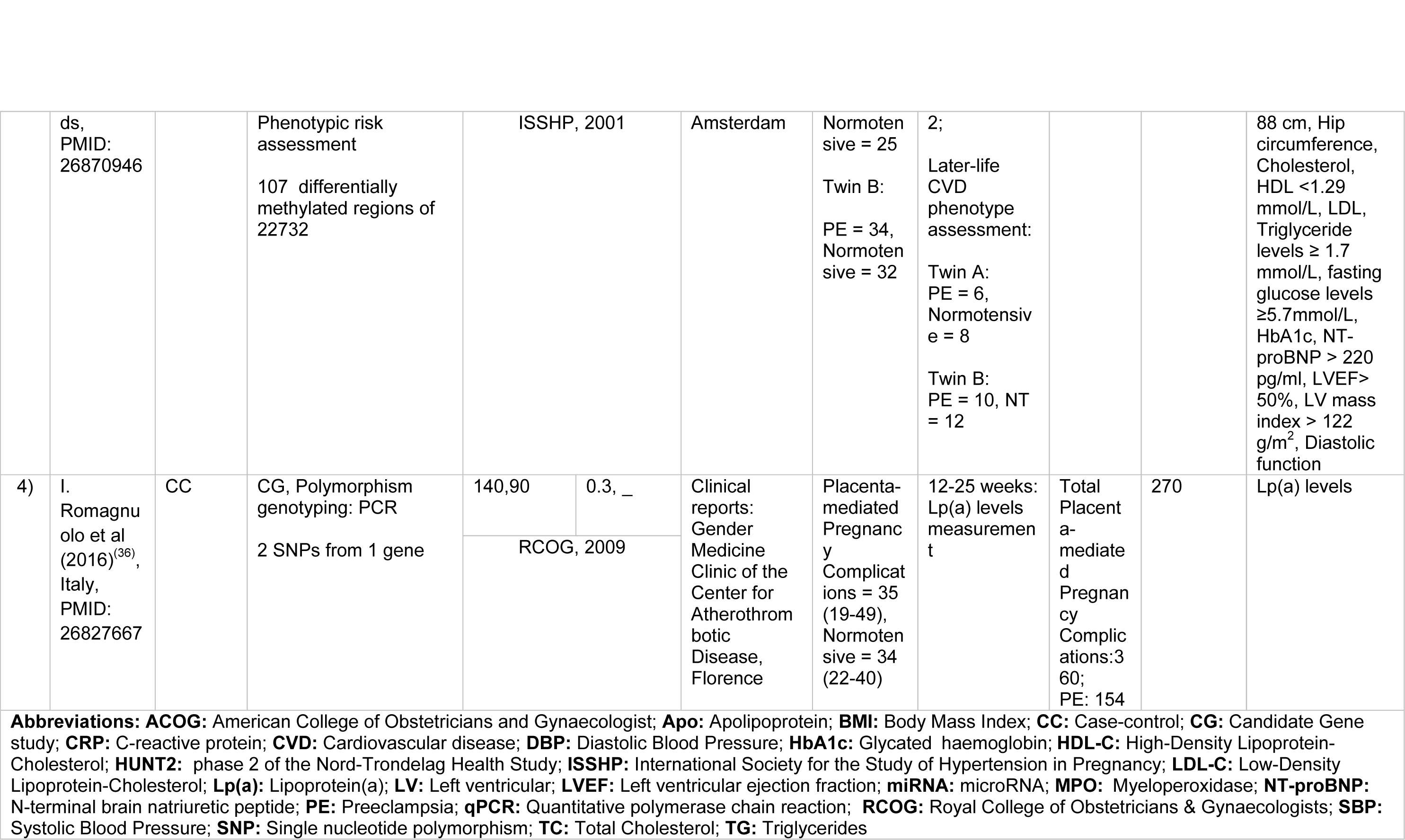
General Characteristics of studies on PE and CVD risk factors.

The study populations were primarily of European ancestry, with the total number of participants ranging from 4 to 2945 and maternal ages ranging from 27 to 49.5 years. The follow-up time after the index pregnancy varied from 12 weeks to 22 years. The International Society for the Study of Hypertension in Pregnancy^(42)^, the American College of Obstetricians and Gynaecologists^(43)^ and the Royal College of Obstetricians and Gynaecologists’^(44, 45)^ diagnostic criteria were used in four studies to identify women with PE. The CVD outcome measures included premature acute coronary syndrome (ACS)^(38, 39)^, angina pectoris^(40)^, myocardial infarction^(40)^, cerebral stroke^(40)^ and cardiometabolic risk traits such as blood pressure^(35, 37, 40)^, glucose levels^(35, 37, 40)^, weight^(37)^, BMI^(35, 40)^, waist circumference^(35, 37)^, hip circumference^(35, 37)^, left ventricular ejection fraction (LVEF)^(35)^, left ventricular mass index^(35)^, diastolic function^(35)^, apolipoprotein A (Apo A)^(37)^, apolipoprotein B (Apo B)^(37)^, low-density lipoprotein-cholesterol (LDL-C)^(35, 37)^, high-density lipoprotein-cholesterol (HDL-C)^(35, 37, 40)^, total cholesterol^(35, 37, 40)^, triglyceride levels^(35)^ and lipoprotein (a) [Lp(a)] levels^(36)^.

### CVD endpoints

Two studies were included on the progression of PE to CVD endpoints (Table 1 and 3)^(38, 39)^. Both studies focused on circulating miRNAs, their association with PE, and premature ACS, the outcome of the studies^(38, 39)^.

In the first study, Dayan et al. investigated 372 miRNAs in women who developed premature ACS 14.2 years after PE (N=30) versus those who had normotensive pregnancies (N=146). This led to the identification of 16 differentially expressed miRNAs in PE, which consisted of: (i) an increase of 10 miRNAs with a fold change of 1.3-2.0; and (ii) a decrease of 6 miRNAs with a fold change of 1.3-2.8. However, of the 16 miRNAs, only three miRNAs that met the eligibility requirements for evaluations in larger validation cohorts were selected in the study. Moreover, the three chosen miRNAs were also associated with various biological mechanisms involved in CVD risk^(39, 46)^. The mir-1225p was previously associated with hepatic lipid metabolism, miR-126-3p with angiogenesis and miR-146a-5p with anti-inflammation^(39, 46)^. However, in this study, all three miRNAs (miR-122-5; miR-126-3p; and miR-146a-5p) significantly lowered in women with premature ACS and a history of PE, even after adjusted for chronic hypertension^(39)^ (See Table 1 and 3 for more details).

In the second study, 2578 miRNAs were screened comparing circulating miRNA levels between four cohorts: (i) an ACS cohort with a history of PE (N= 18) versus normotensive (N=17); (ii) a non-ACS cohort with a history of PE (N= 20) versus normotensive (N=20); (iii) an ACS versus non-ACS cohort; and (iv) women with PE versus normotensive without ACS^(38)^. The development of premature ACS varied from 16- and 19-years post PE across the ACS and non-ACS cohorts respectively. Among the 2578 miRNAs screened, only one miRNA (miR-206) was altered in all four cohorts. However, a history of PE was linked to approximately ten-fold lower plasma levels of miR-206 in women with ACS compared to a history of normotensive pregnancy. This was confirmed in a second cohort of women without ACS, but the change was more moderate at 1.8-fold. Moreover, through miRNA pathway enrichment analysis, Wnt-signalling was identified as the most significantly modified pathway common to PE and ACS. Besides, the most interacting genes with miR-206 in the gene target interaction network were identified as Nuclear Factor of Activated T-cells 5 (*NFAT5)*, Cyclin D2 *(CCND2)* and Mothers Against Decapentaplegic homolog 2 (*SMAD2)*^(38)^ (See Table 1 and 3 for more details).

### CVD risk factors

Four case-control studies were included that investigated CVD risk factors (Tables 2 and 3)^(35–37, 40)^. Two were candidate gene studies on PE and later-life hypertension^(37, 40)^. The later-life hypertension was considered as the outcome in both studies.

**Table 3:** Results of Included Studies.

| Table 3: Results of Included Studies |  |  |  |  |  |
| --- | --- | --- | --- | --- | --- |
| SI no: | Author, Year | Gene names/<br>miRNA's | SNPs | Statistical analyses | Effect Size; *P*-value<sup>∞</sup> |
| Candidate Gene Studies |  |  |  |  |  |
| 1) | A.S. Kvehaugen, 2014<sup>(40)</sup> | *RGS2* | C1114G: rs4606 | Logistic regression,<br>Hosmer-Lemeshow<br>goodness of fit | Later- life hypertension and Early-Onset PE:<br>**CG:** OR = 7.90, 95% CI (1.30-4.82)<sup>a</sup>; 0.025<br>**CG+GG:** OR= 7.96, 95% CI (1.33-47.8)<sup>a</sup>; 0.023<br>Later-life hypertension and PE:<br>**GG:** OR = 1.46, 95% CI (1.02-2.09)<sup>b</sup>; 0.037<br>Stage 2 hypertension and PE:<br>**GG:** OR = 1.93, 95% CI (1.05-3.53); 0.033<br>**CG+GG:** OR = 1.43, 95% CI (1.02- 2.00); 0.036 |
| 2) | I. Romagnuolo, 2016<sup>(36)</sup> | *LPA* | +93C>T: rs1853021,<br>+121G>A: rs1800769 | Chi-square test,<br>Kruskal-Wallis test,<br>linear regression | As allelic burden ↑<br>Lipoprotein(a) concentrations ↑<br>*P* = 0.001<br>Influence of allelic burden on PE and still birth risk<br>*P* = 0.06 |
| 3) | I.V. da Silva, 2021<sup>(37)</sup> | *AQP3* | rs2231231 | Fisher exact test, Chi-Square, binary logistic regression, dominant and recessive models | Later-life hypertension:<br>**AA+AC:** OR = 3.53, 95% CI (1.24-10.04)<sup>d</sup>; 0.018<br>**AA:** OR = 7.22, 95% CI (1.59-32.82)<sup>d</sup>; 0.011<br>**AQP3** (rs2231231) correlated with ↑ CVD risk in women with prior PE<br>**AA+AC:** Genotype distributions different in the 4 groups of women<sup>e</sup>; *P* = 0.026 |
| **MicroRNA Studies** |  |  |  |  |  |
| 4) | N. Dayan, 2018<sup>(39)</sup> | miR-122-5p, miR-126-3p, miR-146a-5p | — | Multivariate linear and logistic regression, Spearman correlation coefficients | History of PE in ACS patients:<br>**miR-122-5p:** OR = 0.66, 95% CI (0.47 - 0.92)<sup>c*</sup>; 0.009<br>**miR-126-3p:** OR = 0.48, 95% CI (0.29 - 0.78)<sup>c*</sup>; 0.002<br>**miR-146a-5p:** OR= 0.57, 95% CI (0.35 - 0.91)<sup>c*</sup>; 0.017 |
| 5) | K. Schlosser, 2020<sup>(38)</sup> | miR-206 | — | D'Agostino Pearson | **miR-206** altered in all 4 cohorts. |
|  |  | Most significant altered enrichment pathway common to PE and ACS: Wnt signaling<br>Most interacted genes with miR-206 in the gene target interaction network: *NFAT5*, *CCND2*, *SMAD2* |  | test, Mann-Whitney or unpaired t-test, Fishers Exact test, *P*-values and Benjamini-Hochberg false discovery rate (FDR) values calculated using EdgeR statistical software package | ACS Cohort 1: (prior PE Vs normotensive)<br>Fold change = -10.6; FDR adjusted *P*-value<sup>#</sup> = 6.98E-04<br>Non-ACS Cohort 2: (prior PE Vs normotensive)<br>Fold change = -1.8; FDR adjusted *P*-value<sup>#</sup> = 6.21E-01<br>Cohort 3: (ACS Vs Non-ACS)<br>Fold change = 6.9; FDR adjusted *P*-value<sup>#</sup> = 8.65E-09<br>Cohort 4: (Current PE Vs normotensive: published literature)<br>Fold change = 1.4, miR-206: upregulated in PE |
| Epigenetic Study |  |  |  |  |  |
| 6) | C. Oudejans, 2016<sup>(35)</sup> | Genes associated with 12 differentially methylated regions with potential CVD risk following PE:<br>*AK056657, HIVEP3, PCDHA1, STAG2, NPIPL1, SRPK3, PSMD4, C4orf48, AB020652, DHX58, IGHE* | — | Statistical testing for Differential DNA methylation, statistical correction for multiple hypothesis testing and ranking conducted using SeqMonk tool,<br>Statistical filter> Chi-square> fwd/rev | Effect size not mentioned |
| **Note:** ∞: *P* significance < 0.05; *: miRNAs inversely associated with history of PE; #: FDR-adjusted *P*-value <0.05<br>**a:** adjusted for age and BMI; **b:** adjusted for history of PE; **c:** adjusted for chronic hypertension; **d:** adjusted for age; **e:** Group 1: PE and hypertensive women, Group 2: PE and normal blood pressure post-pregnancy, Group 3: normal blood pressure during pregnancy and hypertensive women, Group 4: normal blood pressure during and post-pregnancy<br>**Abbreviations:** **ACS:** Acute Coronary Syndrome; **95% CI:** 95% Confidence Interval; **CVD:** Cardiovascular disease; **miRNAs:** micro RNAs; **OR:** Odds Ratio; **PE:** Preeclampsia; **SNPs:** Single Nucleotide Polymorphisms |  |  |  |  |  |

In the first study, four genetic variants from four genes: Aquaporin-3 (*AQP3*; rs2231231), Aquaporin-7 (*AQP7*; rs2989924), Nitric oxide synthase 3 (*NOS3*; 4B/A intron) and Cytochrome B-245 Alpha Chain (*CYBA*; rs4673) were investigated for their association with later-life hypertension, in a cohort of women with prior PE (N=48) or who had a previous normotensive pregnancy (N= 98)^(37)^. Previous studies have identified that all four genes included in the analysis play an essential role in redox homeostasis and oxidative stress, which are major components of a metabolic syndrome (a group of risk factors specific to CVD)^(37, 47, 48)^. However, among the four SNPs, only one SNP rs2231231 from the *AQP3* gene, dominant and recessive model of A allele [AA+AC] and [AA] respectively, was associated with PE and the development of hypertension in women 2–16 years post-pregnancy. Moreover, in the study, the *AQP3* (rs2231231), [AA+AC] was also linked to a greater risk of CVD in women with a history of PE, as it showed different genotype distributions in four different groups of women. Group 1 included preeclamptic and hypertensive women, group 2 included preeclamptic women and those with normal blood pressure post-pregnancy, group 3 included women with normal blood pressure during pregnancy and hypertensive women, and group 4 included women with normal blood pressure during and post-pregnancy^(37)^ (See Table 2 and 3 for more details).

The second study investigated a single genetic variant (rs4606) in the Regulator of G Protein Signaling 2 (*RGS2*) gene to check its association with later-life hypertension 15-years following PE. This analysis was conducted in Norwegian PE cases (N= 934) and controls (N= 2011)^(40)^. Past studies identified that numerous vasoconstrictors are negatively regulated by the regulator of G protein signaling 2^(49)^. Moreover, CG or GG genotypes of rs4606 in the *RGS2* gene have been previously found to be associated with PE women^(50)^. However, this study identified the association of rs4606 [CG, CG+GG] polymorphism with the risk of later-life hypertension and a history of early-onset PE after adjusting for age and BMI. In addition, associations were also identified with the SNP, rs4606 [GG], later-life hypertension and a history of PE and between rs4606[GG, CG+GG], later-life stage 2 hypertension and a history of PE^(40)^ (See Table 2 and 3 for more details).

The third CVD risk factor study is a candidate gene study on PE and Lp(a) levels^(36)^. The Lp(a) levels were considered the study’s outcome. Lp(a), which is associated with the plasminogen-like glycoprotein, is a significant risk factor for atherosclerotic CVD, mainly in those with LDL-C or HDL-C^(51, 52)^. Moreover, from previous studies, Lp(a) levels were also observed to be increased in women with a history of PE^(53, 54)^. However, this research was conducted in a cohort with a history of various placenta-mediated pregnancy complications (N= 360), including PE (N= 154), stillbirth (N= 121), and small-for-gestational-age (N= 85) as cases. Healthy women with no history of vascular disorders (conditions that affects blood vessels, e.g., venous thrombosis, Aneurysm) and pregnancy complications were included as control groups. The study investigated the involvement of the two polymorphisms (rs1853021: +93C > T and rs1800769: 121G > A) in the Lipoprotein A (*LPA)* gene in modifying Lp(a) levels and placenta-mediated pregnancy complications risk. The Lp(a) levels were analysed 12 weeks post-pregnancy, and it was observed that women with a history of PE and stillbirth had elevated Lp(a) levels. Moreover, as the unfavourable allelic burden of *LPA* gene elevated, Lp(a) concentrations gradually increased. A similar association of increased risk with PE and stillbirth with Lp(a) levels was identified, although it was not significant (*P*=0.06) (See Tables 2 and 3 for more details).

The last included article used whole-genome bisulphite DNA methylation study with two identical twin sister pairs discordant for PE^(35)^. This study investigated the epigenomic alterations associated with CVD risk following PE. The twin sister pairs were examined for epigenetic risk two years post-pregnancy. Moreover, 18 CVD markers were tested between the twin sisters to understand the phenotypic risk 6-12 years post-pregnancy; no differences were observed. However, a genome-wide methylC-sequencing of 22732 differentially methylated regions (DMRs) revealed 107 DMRs significantly altered in all individuals. Among these, 12 DMRs were found to be shared by the affected twin sisters, with at least half a difference in their methylation percentage and having the same up or down-regulation. These findings were vastly different from those of their unaffected twins (Table 3). These 12 DMRs may be potentially linked to CVD risk following PE, and the genes associated with the regions can be found in Table 3. One of these genes *DHX58* was found to be associated with coronary artery disease in another study^(55)^. However, the remaining genes associated with DMRs were linked to CVD mainly in animal studies^(56, 57)^. The authors of this epigenetic study concluded that the ongoing long-term CVD risk in the affected twin sister might be due to the changes in her DNA methylation caused by PE.

## DISCUSSION

### Main findings

In this systematic review, we surveyed the published literature to identify common evidence on shared genomic factors associated with CVD endpoints or risk factors following PE available as of February 2^nd^,2023. Following quality control and screening, we identified six case-control studies longitudinally testing for CVD endpoints or CVD risk factors following PE^(35–40)^. All the included studies were of case-control design and European ancestry. Both manuscripts investigating CVD endpoints focussed on premature ACS following PE using miRNA markers^(38, 39)^. These studies identified four miRNAs (miR-122-5p, −126-3p, −146a-5p, −206) differentially expressed in women with premature ACS following PE and concluded that these findings might offer better insights into biological mechanisms that could be responsible for the elevated risk of CVD post-PE^(38, 39)^.

The CVD risk factors category consisted of: i) two candidate gene studies^(37, 40)^ on PE and later-life hypertension ii) one candidate gene study^(36)^ on PE and Lp(a) levels and iii) an epigenetic study on PE and later-life CVD risk factors^(35)^. The first candidate gene study identified one SNP rs2231231 from the *AQP3* gene associated with PE and later-life hypertension^(37)^. The study concluded that as the *AQP3* gene was only associated with hypertension post-pregnancy, the role of the gene might be linked to later-life hypertension risk factors including oxidative stress and metabolic syndrome^(37)^. The second candidate gene study also identified another SNP rs4606 from the *RGS2* gene associated with PE and later-life hypertension^(40)^. In this analysis, even after accounting for rs4606 SNP and other CVD risk factors, PE continued to be a standalone risk factor for future hypertension^(40)^. Moreover, from another candidate gene study on PE and lipoprotein(a) [Lp(a)] levels, two polymorphisms (rs1853021: +93C > T and rs1800769: 121G > A) in the *LPA* gene were observed in modifying Lp(a) levels and placenta-mediated pregnancy complications risk^(36)^. The study detected that, those women with a history of PE and stillbirth had an increased concentration of Lp(a). This research helped in confirming the relationship between pregnancy complications and the atherothrombotic marker, Lp(a)^(36)^. Finally, from the epigenetic study, 12 DMRs associated with CVD risk following PE were identified^(35)^. The study concluded that the whole-genome bisulfite DNA methylation sequencing approach used in this study would help in identifying biomarkers that can be used for early CVD risk stratification for women after a complicated pregnancy^(35)^.

None of the included studies reached the maximum score on the Newcastle-Ottawa Scale, as most studies did not include their non-response rate (Tables S1-S6). Thus, the non-response bias that may have existed in the studies could not be calculated. The limited amount of primary research demonstrates a critical gap in the literature that needs to be addressed. Hence, despite the increasing number of publications on PE and its relationship with later-life CVD^(18, 19, 58–60)^, more empirical research is required to identify the genomic factors involved in the progression to CVD after a history of PE using a longitudinal study framework.

### Interpretation

Several systematic reviews and meta-analyses discuss CVD risk following PE^(61–64)^, but only a small number consider the shared genomic risk factors associated with both diseases^(65, 66)^. In addition, many of these previous systematic reviews make conclusions combining the results between animal and human studies^(66)^. While animal models on PE are well established across different species, the focus on CVD *in vivo* models is heterogeneous in nature and does not always capture the complexity of how biological processes may evolve. Hence, a standard animal model does not work for CVD, and several animal models or a personalised model would be required for a better understanding, increasing the complexity^(67)^. Thus, this systematic review focused on including studies on genetic, epigenetic or transcriptomic factors associated with women with a history of PE and later-life CVD risk.

### Strengths

A comprehensive analysis was conducted to systematically identify up-to-date studies on genomic loci associated with the progression of PE to CVD. Only studies conducted using the same cohorts of women with CVD and PE, and articles on CVD risk following PE, were included. This would help better identify the overlapping genomic risk loci of both diseases over time without any bias. The scores were given based on the checklist relevant to their study design. Moreover, the criteria for ascertainment of exposure in the Newcastle-Ottawa scale were tailored to incorporate genetic or epigenetic assessments. These customizations in quality assessments have helped thoroughly analyse the studies, irrespective of their study type or design.

### Limitations

This study has certain drawbacks that need to be addressed. First, each included literature had different methodologies, study designs, findings, and analyses of the CVD outcomes. Thus, a meta-analysis could not be performed because of the significant heterogeneity between studies. Second, the sample size of each included study was comparatively small. Third, only articles published in English were included. Fourth, as all included studies were of European ancestry, there was no data relevant to other ethnicities.

## RECOMMENDATION

Identifying the genomic risk loci associated with the progression of PE to CVD would provide a better understanding of the underlying biology of both diseases. Therefore, more comprehensive longitudinal research is required directed to this aim. Moreover, the direction of causality needs to be determined using Mendelian randomisation. Also, the self-reported surveys need to be further validated using electronic health records and study populations with larger sample sizes should be included.

## CONCLUSION

We conducted an extensive systematic review of the literature that demonstrates limited publications regarding genomic risk loci associated with the progression of PE to later-life CVD. This review provides critical insight into the heterogeneous nature of genomic studies investigating CVD following PE and highlights the urgent need for large scale longitudinal studies that investigate the genetic risk underlying the progression to CVD following PE.

## Data Availability

This is a systematic review and all data and results are contained within this manuscript.

## Registration and protocol

This systematic review was not registered in any publicly available registry.

## Contribution to authorship

P.M, E.M, and S.B conceived the project idea, while G.K and H.P designed the analysis workflow. G.K, T.N, and N.T screened all the relevant articles and performed quality assessment analysis. G.K wrote the manuscript, with supervision from P.M. All authors contributed to the final manuscript.

## Acknowledgements

We would like to acknowledge National Health and Medical Research Council (NHMRC), Australia, research grant and the Tasmania Graduate Research Scholarship (TGRS) for their generous financial support of this research. We are also grateful to Menzies Institute for Medical Research, the University of Tasmania for providing access to the research facilities and resources.

## Details of Ethical approval

Ethical approval of the institutional review board was not required.

## List of Abbreviations

PE: Preeclampsia
CVD: Cardiovascular disease
PRISMA: Preferred Reporting Items for Systematic Reviews and Meta-Analyses
BMI: Body mass index
*LCT*: Lactase
*LRP1B*: Low density lipoprotein receptor-related protein 1B
*GCA*: Grancalcin
*RND3*: Rho Family GTPase 3
DNA: Deoxyribonucleic acid
miRNA: MicroRNA
HELPP: Hemolysis, Elevated Liver enzymes and Low Platelets
ACS: Acute coronary syndrome
LVEF: Left ventricular ejection fraction
Apo A: Apolipoprotein A
Apo B: Apolipoprotein B
LDL-C: Low-density lipoprotein cholesterol
HDL-C: High-density lipoprotein cholesterol
Lp(a): Lipoprotein(a)
*NFAT5*: Nuclear Factor of Activated T-cells 5
*CCND2*: Cyclin D2
*SMAD2*: Mothers Against Decapentaplegic homolog 2
*AQP3*: Aquaporin-3
*AQP7*: Aquaporin-7
*NOS3*: Nitric oxide synthase 3
*CYBA*: Cytochrome B-245 Alpha Chain
*RGS2*: Regulator of G Protein Signaling 2
DMR: Differentially methylated regions
*DHX58*: DExH-Box Helicase 58
*LPA*: Lipoprotein(A)

## Supplemental Material

PRISMA 2020 CHECKLIST

SEARCH STRATEGY: (27/08/2021)

UPDATED SEARCH: (02/02/23)

TABLES S1-S6

